# Autonomic dysfunction after moderate-severe traumatic brain injury: symptom spectrum and clinical testing outcomes

**DOI:** 10.1101/2021.06.29.21259552

**Authors:** Lucia M. Li, Ekawat Vichayanrat, Martina del Giovane, Helen HL Lai, Valeria Iodice

## Abstract

**Background and Objective:** Survivors of moderate-severe traumatic brain injury (msTBI) frequently experience chronic, debilitating somatic symptoms, which are largely unexplained. The phenomenon of paroxysmal sympathetic hyperactivity, reflecting hyperacute autonomic dysfunction, is well-documented after msTBI. Limited animal and human studies, using experimental measures, have found evidence for autonomic dysfunction after msTBI. However, no studies have investigated the range and type of autonomic symptoms and autonomic dysfunction existing in msTBI.

We set out to investigate the presence and type of subjective and objective autonomic dysfunction in msTBI.

**Methods:** We conducted two cohort studies. Cohort 1 comprises msTBI patients prospectively recruited from a national TBI outpatient clinic, in whom we assessed burden of autonomic symptoms using the Composite Autonomic Symptom Score (COMPASS31) autonomic symptom questionnaire. Cohort 2 comprises msTBI patients who had standard clinical autonomic function testing (supine/tilted catecholamine levels, head-up tilt, Valsalva manoeuvre, respiratory sinus arrhythmia assessment), retrospectively identified from the database of a regional clinical autonomic unit.

**Results:** Cohort 1 comprises 29 msTBI patients (6 females, median age 40 years, range 19-76), with a median time since injury of 19 months (range 4-105). There was multi-domain symptom burden, with all but 3 patients reporting symptoms on the COMPASS31 questionnaire, and 17/29 reporting symptoms in 3+ domains. The most commonly reported symptoms were gastrointestinal (22/29), followed by orthostatic (17/29), pupillomotor (14/29), secretomotor (14/29), bladder (12/29) and, least commonly, vasomotor (6/29). Cohort 2 comprises 18 msTBI patients (7 females, median age 44 years, range 21-64), with a median time between injury and testing of 57.5 months (range: 2-416). The majority of patients (15/18) had orthostatic symptoms as part of the reason for referral. Clinical autonomic function testing revealed a broad spectrum of autonomic dysfunction: 3/18 had evidence of sympathetic dysfunction, 10/18 had evidence of parasympathetic dysfunction, of which 6 also had evidence of mixed dysfunction.

**Discussion:** Our results provide evidence for clinically relevant autonomic dysfunction after moderate-severe TBI at the chronic stage. Our study advocates for routine enquiry about potential autonomic symptoms in this population, and the utility of formal clinical autonomic testing in providing diagnoses.

## INTRODUCTION

It is increasingly recognised that survivors of Traumatic Brain Injury (TBI), particularly more severe injuries, are left with high rates of chronic neurologic, cognitive and behavioural sequelae. What is less recognised in the clinical community is that TBI survivors often also report long-lasting systemic symptoms, which can be just as debilitating. Many of these symptoms are suggestive of autonomic nervous system dysfunction^12^. Changes in heart rate variability, an experimentally used measure of cardiovascular autonomic nervous system (ANS) function, has been found in a number of studies of concussion and mild TBI, with one study reporting a positive correlation between TBI severity and degree of both sympathetic parasympathetic impairment^3–6^. Paroxysmal sympathetic hyperactivity, manifesting as episodic surges in heart rate and blood pressure, temperature, sweating, respiratory rate, sometimes associated with abnormal posturing, mostly frequently observed in acute cases of severe TBI, is thought to represent autonomic dysfunction and relates to worse clinical outcomes^7^.

Orthostatic symptoms, pre-syncopal and syncopal episodes are those most likely to be noticed and recognised as being autonomically-mediated. However, ANS dysfunction can manifest in a range of additional different symptoms which have the potential to cause significant negative impact on daily function and quality of life, such as abnormal sweating, nausea and bloating, constipation and diarrhoea, dry eyes and mouth, changes in skin colour and thermodysregulation, erectile dysfunction in men, visual blurring and photosensitivity^8^. Symptoms of autonomic dysfunction can be quantitatively described with the COMPASS-31 tool, a simple-to-administer questionnaire that captures a range of symptoms which are sensitive and specific to autonomic dysfunction^9^. Clinical autonomic function tests seek to non-invasively assess the integrity of the ANS across multiple modalities, including cardiovagal, sudomotor and adrenergic function, and have shown utility in characterising and tracking ANS dysfunction^10^.

To date, no study has specifically investigated autonomic function after TBI. We present the results of a clinical investigation in two cohorts of chronic, severe TBI patients. First, we present clinical autonomic function testing results from a cohort retrospectively identified from referrals to a national referral Autonomic centre, in order to show the characteristics of autonomic dysfunction in chronic severe TBI. Secondly, we collected quantitative symptom assessments in a cohort prospectively identified from a national TBI clinic, in order to understand the clinical symptomology in this cohort.

## METHODS

### Study design and participants

We investigated two separate cohorts of patients with (msTBI), in a cross-sectional study. The first, retrospective cohort was from all patients referred for clinical autonomic function tests to the Autonomics Unit at the National Hospital for Neurology and Neurosurgery, UK. Clinical documentation during testing from all those referred between 2010-2020 was screened with the following terms: TBI, injury, trauma* (wild card to bring up e.g. ‘trauma’, ‘traumatic’), accident. We then searched the electronic health records for details about their injury and medical history. Patients were included for analysis based on the following: history consistent with ms TBI as defined by Mayo Classification^11^ and injury preceded the symptoms for which an autonomics referral was made. Patients were excluded if: TBI was sustained in the context of orthostatic symptoms, TBI occurred with spinal injury, TBI noted but inadequate information to determine its severity or timeline relative to symptoms, an antecedent or concurrent vestibular diagnosis (e.g. vestibular migraine, benign paroxysmal postural vertigo) and the only reported symptom was dizziness, and the medical history included other antecedent or concurrent diagnoses which would explain their symptoms (e.g. long-standing diabetes, multiple systems atrophy) (FIGURE1). The second, prospective cohort was from patients referred to an adult TBI clinic at St Mary’s Hospital London, UK from Jan 2021 to June 2021. All newly referred moderate-severe^11^ TBI patients were invited to complete the COMPASS-31 questionnaire as part the clinic’s cognitive and questionnaire assessments protocol, irrespective of whether they reported symptoms suspicious for ANS dysfunction in their main clinic appointment. The study conforms to the Declaration of Helsinki and ethical approval was granted through the local ethics board [National Research Ethics Service (NRES) Committee London – Camberwell St Giles, IRAS ID: 230221].

**FIGURE 1:**
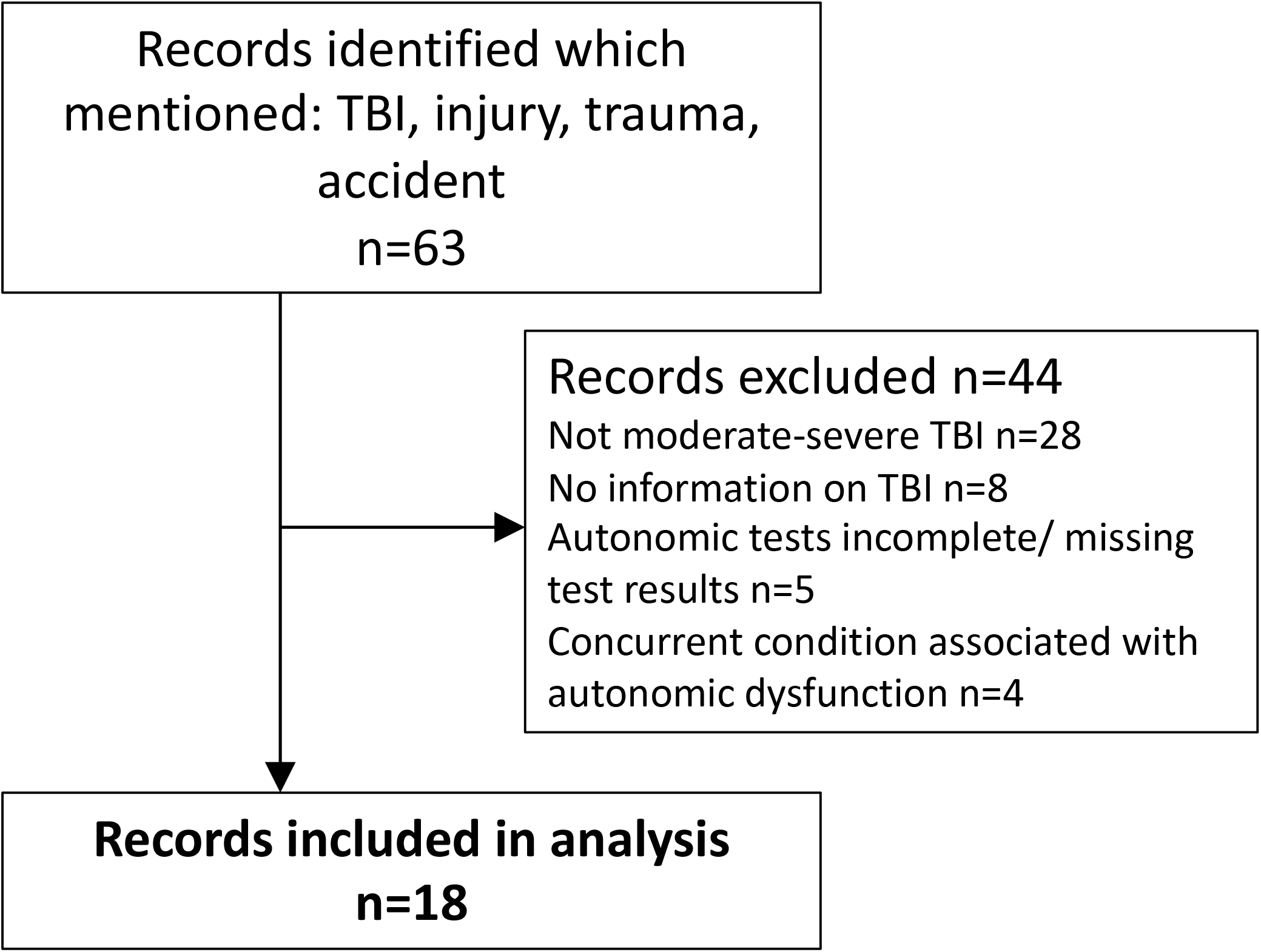
Flowchart denoting records included in retrospective analysis of autonomic testing. TBI=traumatic brain injury

### Autonomic Function Testing: acquisition and analysis

All Autonomic Function Tests (AFTs) were completed at the Autonomics Unit of the National Hospital for Neurology and Neurosurgery between 2010-2020. Before testing, patients are routinely asked to refrain from: heavy exercise for 24 hours, alcohol and caffeine intake for at least 12 hours, and eating for 1 hour. All patients had the standard clinical testing protocol, which included:

- Head up tilt - from supine to 60° head up, and continued for 10 minutes or until patient could no longer tolerate or symptoms of imminent syncope. Beat-to-beat heart rate (HR) and blood pressure (BP) were recorded with a Finapress® or Finometer.
- Supine/ tilted blood noradrenaline levels – used as a biochemical marker of sympathetic neural activity. The plasma level of noradrenaline (NA) was measured using high-performance liquid chromatography.
- Respiratory sinus arrythmia (HR_DB_) – parasympathetic function was assessed by assess the HR response to deep breathing. Patients were asked to breath at a rate of 6 breaths per minute for one minute, and were coached on how long each breath should take. HR_DB_ was the average of the differences between the minimum and maximum heart rates of the 6 breaths. The HR_DB_ was classified as abnormal if it was outside of the age-appropriate range^12^.
- Valsalva test - participants performed a forced expiration, using a 2ml syringe tube, for 15 seconds against a fixed expiratory pressure of 40mmHg for 10-15 seconds. The Valsalva ratio (VR), reflective of both parasympathetic and sympathetic function, is calculated by the maximum HR generated during the Valsalva manoeuvre divided by the lowest HR occurring within 30 seconds of the peak heart rate. The VR was classified as abnormal if it was outside of the age-appropriate range^12^. The BP profile, indicative of adrenergic function, profile was classified as normal/abnormal by the performing clinical scientist, and confirmed by VI and EV.

### Prospective questionnaire symptom assessment

The COMPASS-31 questionnaire was used to assess presence and severity of symptoms which may relate to ANS dysfunction^9^. This questionnaire asks about symptoms in several domains: orthostatic, vasomotor (sweating and skin changes), secretomotor (dry eyes or mouth), gastrointestinal (post-prandial nausea, vomiting or bloating, constipation and diarrhoea), bladder (difficulty passing urine, incontinence) and pupillomotor (photosensitivity and blurred vision). This questionnaire has shown good sensitivity and discrimination value for detecting autonomically-mediated symptoms^13–15^. Each domain is scored separately and then multiplied by a weighting factor, before combining to make a total maximum score of 100. Patients completed the COMPASS questionnaire via the telephone (with LML, MdG or HHLL) or via an online link. A small number of questionnaires were completed face-to-face (by MdG or HHLL).

### Data Availability

Raw scores from the COMPASS31 questionnaire and anonymised clinical autonomic function testing reports from the retrospective cohort are available on request to the corresponding author.

## RESULTS

### Prospective Symptom Burden Assessment Cohort

A total of 29 patients completed the questionnaire (6F/23M, median age 40 years, range 19-76) (TABLE 1A). The median time since injury was 19 months, (range 4-105). The cohort reported multi-domain symptom burden (FIGURE 2). Only 3 patients did not report any symptoms, with 17 of 29 patients having a weighted total score >20, and 17 of 29 patients reporting symptoms in at least 3 subdomains. The subdomain in which symptoms were most commonly reported was the gastrointestinal subdomain with 22 of 29 patients reporting these symptoms, followed by orthostatic symptoms (17/29 patients), then pupillomotor (14/29 patients), then secretomotor (14/29 patients), then bladder (12/29), and vasomotor symptoms were least common (6/29 patients).

**TABLE 1:**
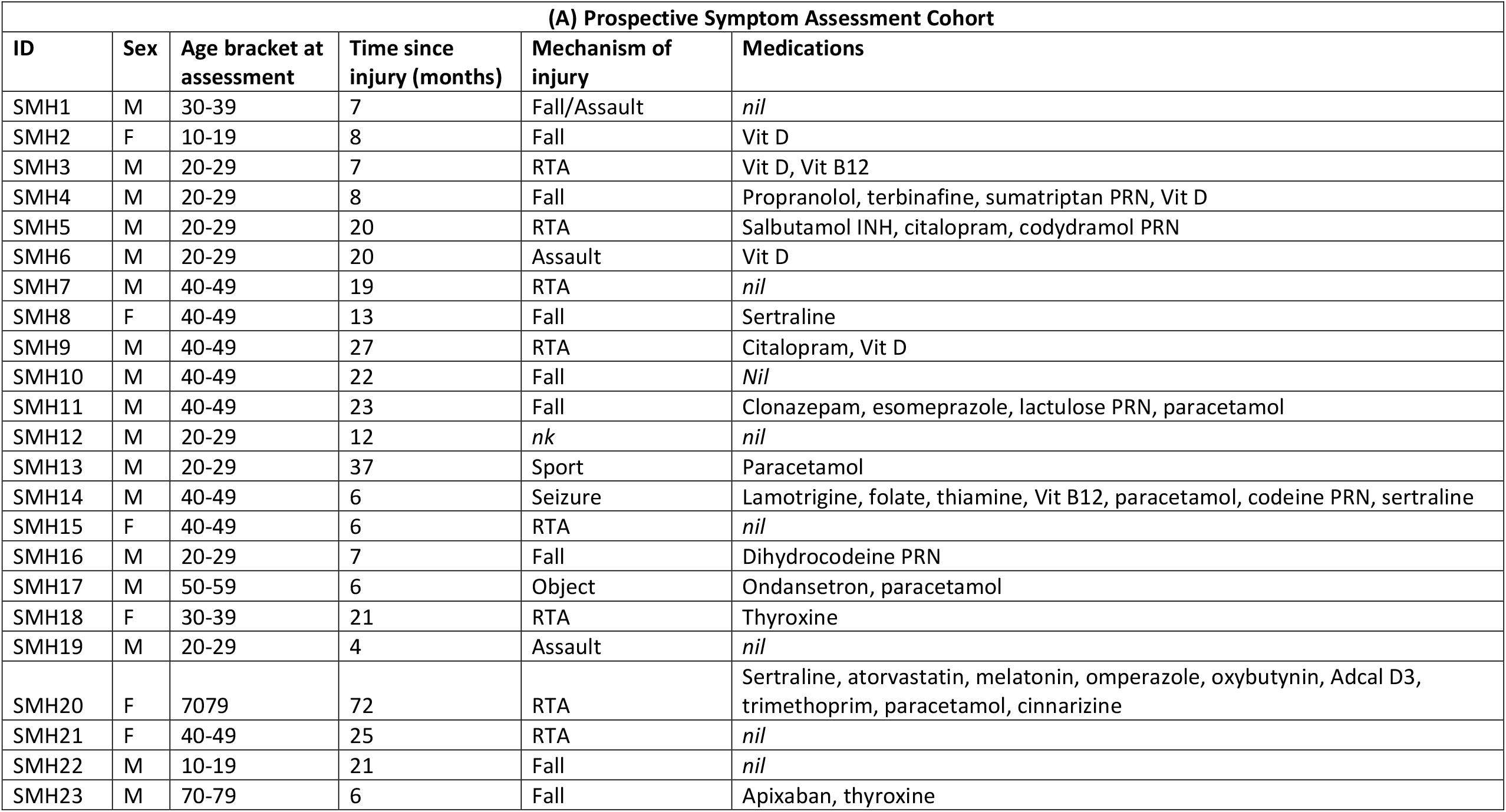
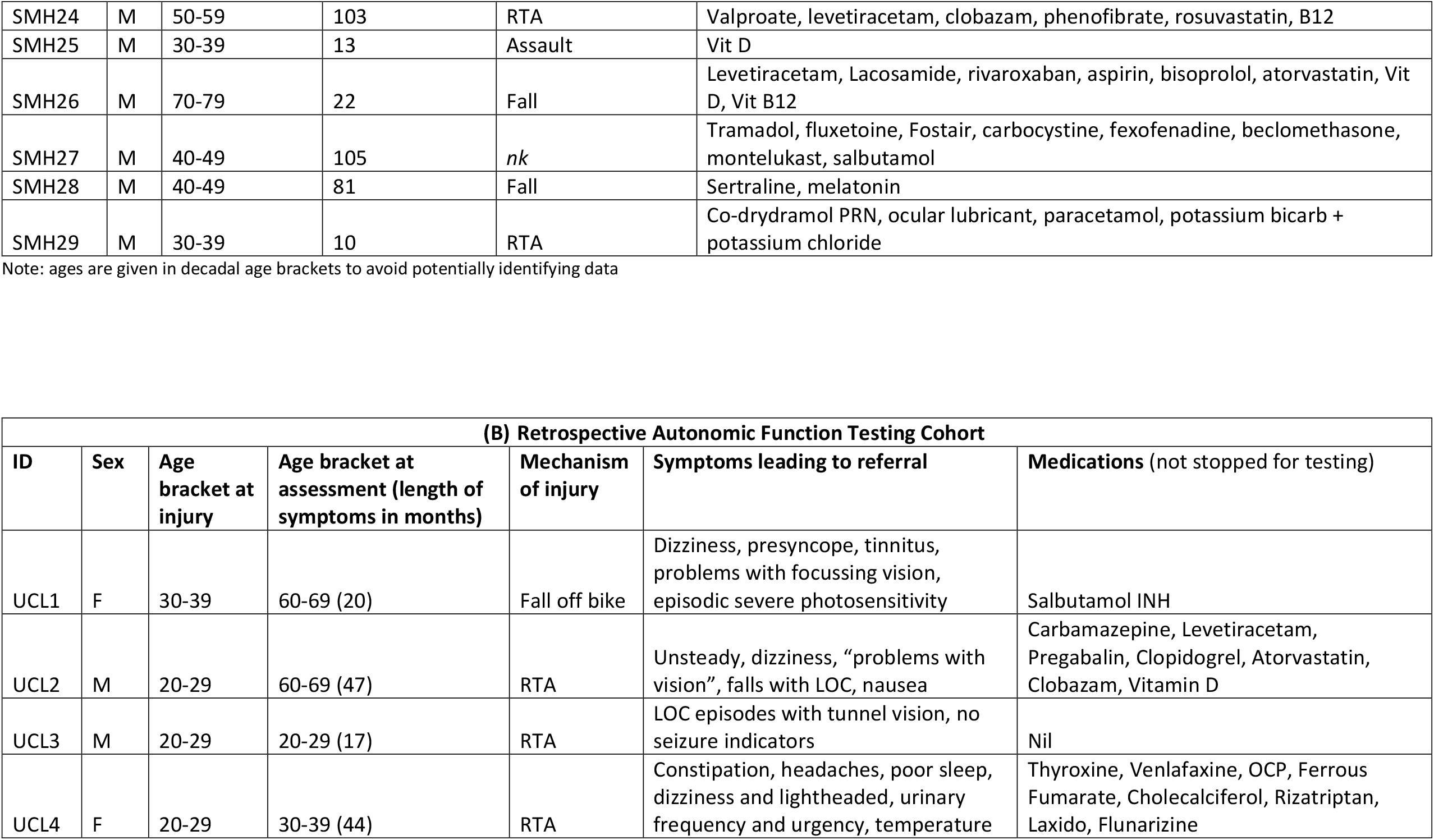
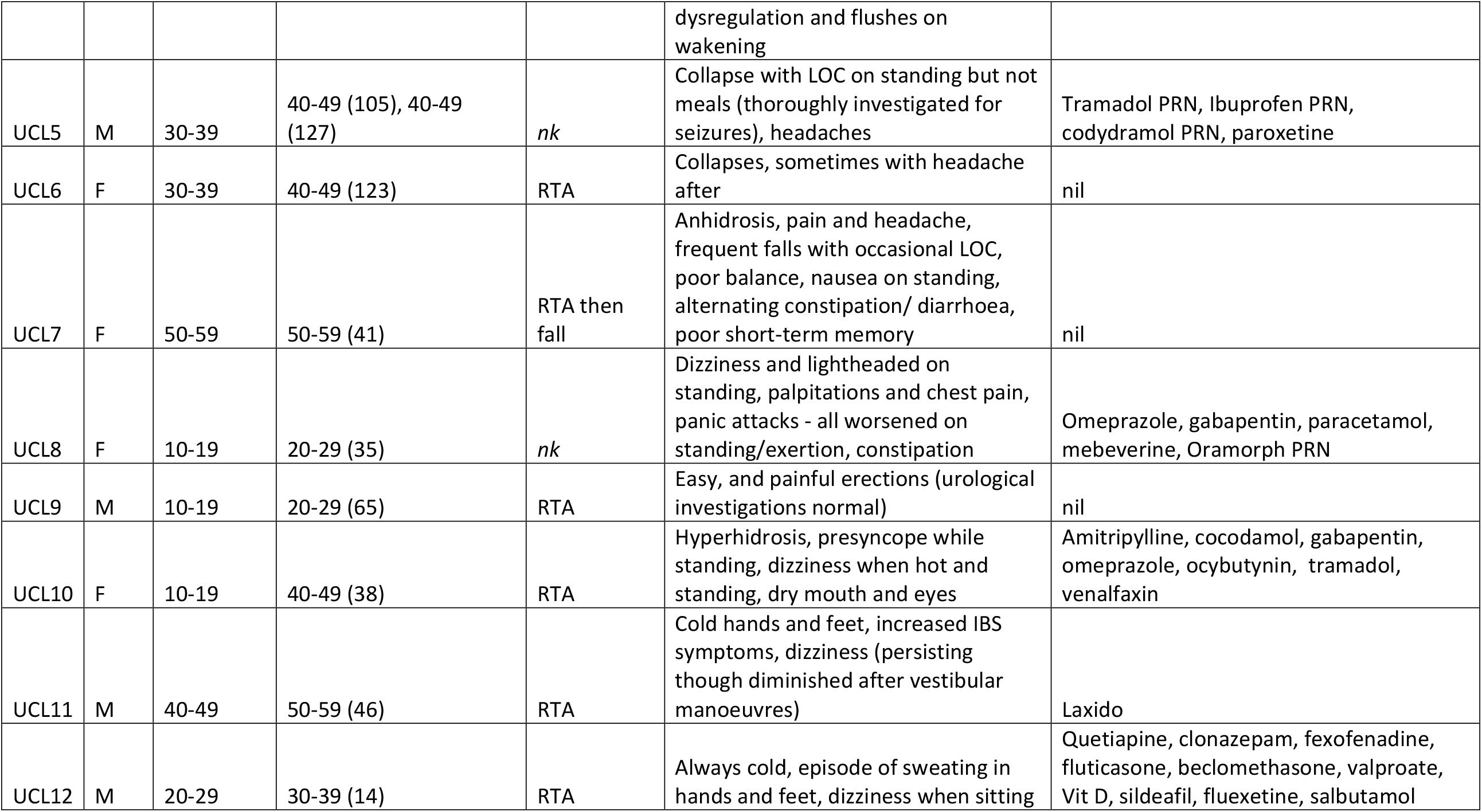
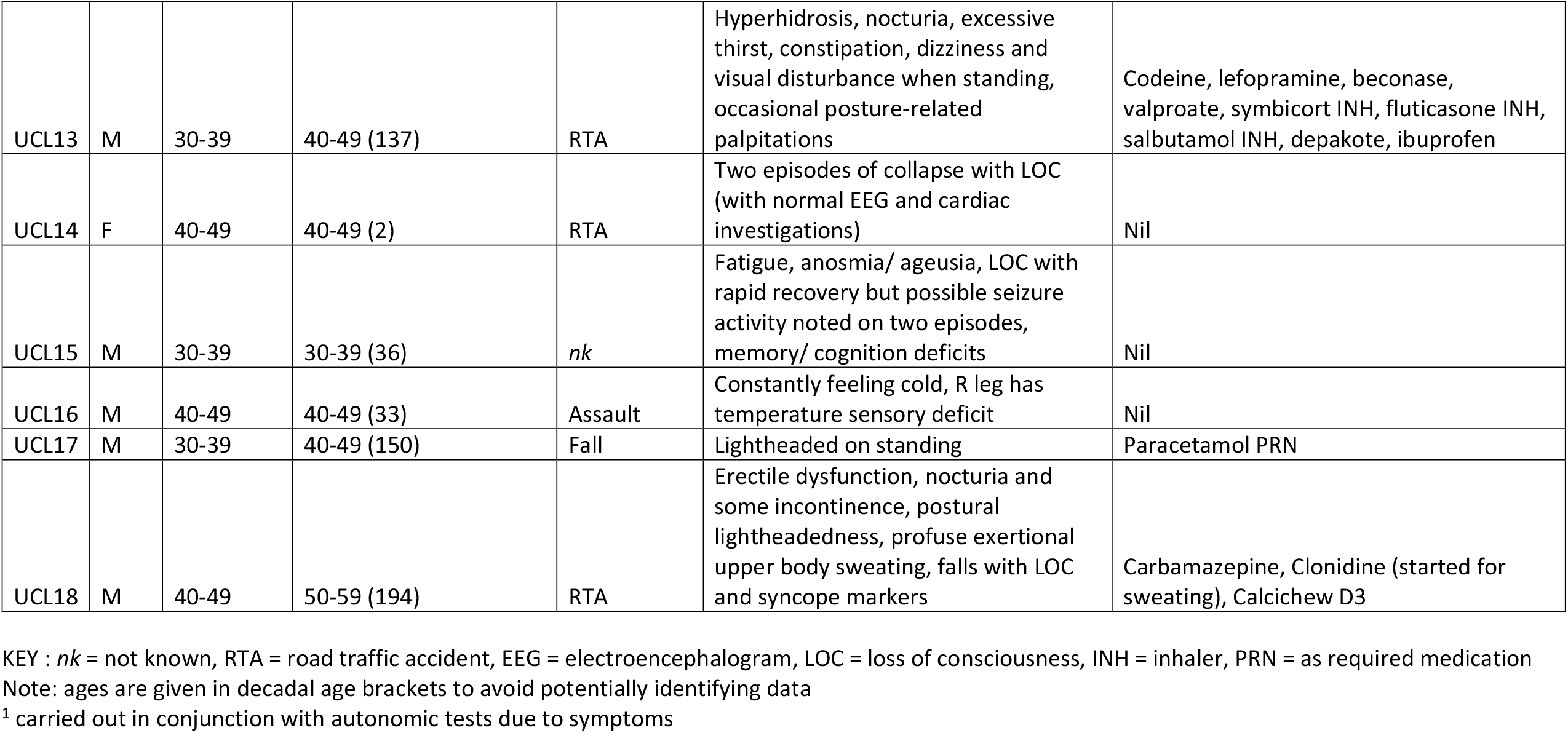
Clinical characteristics of recruited patients. (A) Clinical characteristic of the prospectively recruited symptom assessment cohort (B) Clinical characteristics of the retrospectively recruited autonomic function testing cohort

**FIGURE 2:**
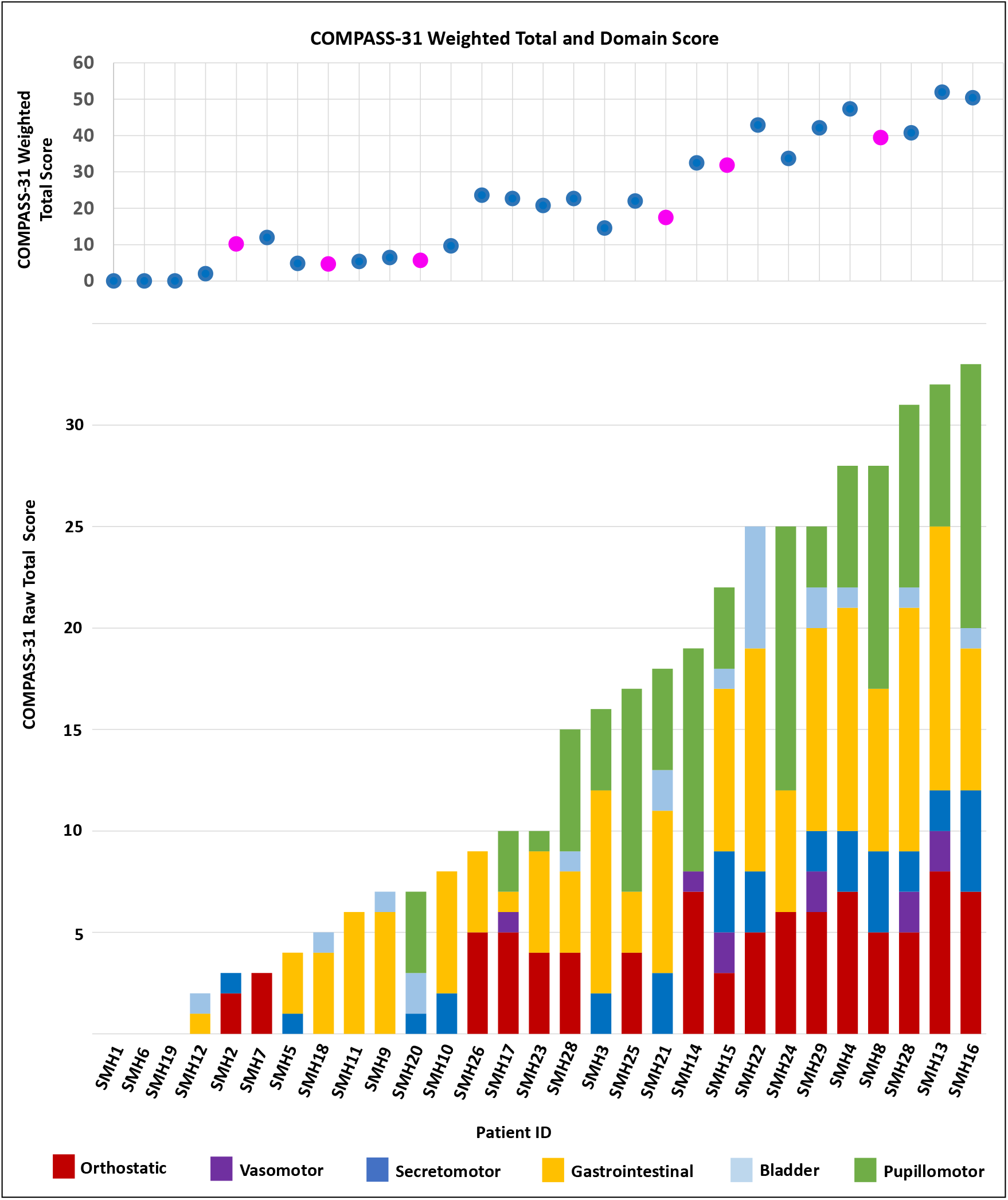
Symptom burden assessed with COMPASS31, with weighted total score for each patient (top panel: each circle represents a patient, pink for female and blue for male patient) and corresponding domain subscore distribution (bottom panel), ordered by raw total score.

### Retrospective Autonomic Function Testing Cohort

A total of 18 patients’ records were identified for further analysis (7F/11M, median age at testing 44 years, range 21-64) (FIGURE1, TABLE 1B). The median time for referral for autonomic investigations was 52.5 months (range: 2-194 months). the median time between the TBI and first testing was 57.5 months (range: 2-416 months); in the majority (12/18) cases, symptoms had first started and were ongoing since their TBI. In this referral cohort, patients were most likely to have reported orthostatic symptoms such as orthostatic intolerance or falls (all but 3 patients reported these) (FIGURE 3A). Changes in sweating was also a prominent reason for referral in 6 of 18 patients. Two patients had been referred as part of work-up for a specific symptom, for which autonomic dysfunction was considered in the differential: patient UCL9 was referred for changes in erectile function, and patient UCL15 was referred for skin and temperature changes affecting a single limb only.

**FIGURE 3:**
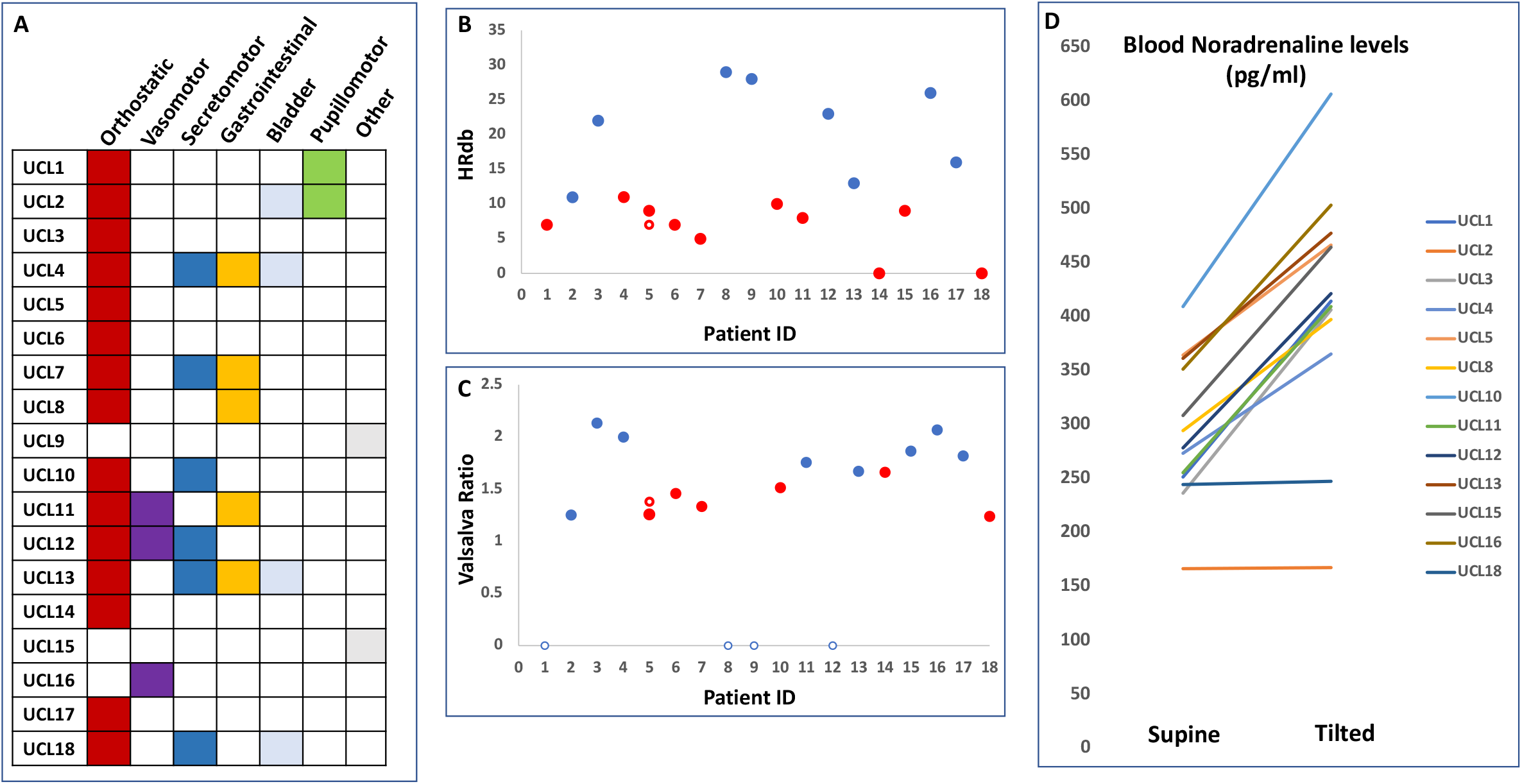
(A) Symptoms reported by each patient, categorized into symptom type, and their subsequent final diagnosis. (B) HR_DB_ for each patient. Where there was no respiratory sinus arrhythmia noted, the value of HR_DB_ is noted as 0. A red dot denotes an abnormal value. The unfilled red circle (UCL5) denotes the value from follow-up testing. (C) Valsalva ratio for each patient. A red dot denotes an abnormal value. The unfilled red circle (UCL5) denotes the value from follow-up testing. An open blue circle denotes Valsava manoeuvre not completed. (D) Supine and tilted blood noradrenaline levels for each patient, where available.

Autonomic Function Testing (AFT) in this cohort demonstrated a broad spectrum of dysfunction (TABLE2). Abnormal parasympathetic function, as denoted by absent or low-for-age respiratory sinus arrhythmia (measured with HR_DB_), was present in 10 of 18 patients (FIGURE 3B). Mixed dysfunction, as denoted by low-for-age Valsalva Ratio (VR), was present in 6 of 18 patients, all of which were included in those with abnormal HR_DB_ (FIGURE 3C). An appropriate rise in blood noradrenaline levels on tilt was absent in 2 patients, suggestive of sympathetic dysfunction (UCL2, UCL18) (FIGURE 3D), one of whom (UCL18) also had abnormal HR_DB_ and VR. An abnormal BP profile on the Valsalva manoeuvre, indicative of adrenergic dysfunction, was noted in UCL2.

**TABLE 2:** Clinical characteristics of recruited patients. Summary of autonomic dysfunction and diagnoses from the autonomic function testing cohort.

| ID | Sympathetic dysfunction | Parasympathetic dysfunction | Mixed dysfunction | Autonomic Diagnoses |
| --- | --- | --- | --- | --- |
| UCL1 |  | X |  |  |
| UCL2 | X |  |  | Orthostatic hypotension |
| UCL3 |  |  |  | Vasovagal syncope |
| UCL4 | X | X |  | Vasovagal syncope, POTS |
| UCL5 |  | X | X | Vasovagal syncope |
| UCL6 |  | X | X |  |
| UCL7 |  | X | X |  |
| UCL8 |  |  |  | Vasovagal syncope |
| UCL9 |  |  |  |  |
| UCL10 |  | X | X |  |
| UCL11 |  | X |  | Vasovagal syncope |
| UCL12 |  |  |  |  |
| UCL13 |  |  |  |  |
| UCL14 |  | X | X |  |
| UCL15 |  | X |  |  |
| UCL16 |  |  |  |  |
| UCL17 |  |  |  |  |
| UCL18 | X | X | X |  |

After head-up tilt testing, 5 patients received diagnoses of vasovagal syncope (UCL3, UCL4, UCL5, UCL8, UCL11) and experienced symptomatic drops in blood pressure on tilt table testing, of which 2 were spontaneous and 3 were provoked by venepuncture or blood removal from an in situ cannula, consistent with reported clinical histories of syncope/pre-syncope. UCL3 experienced a symptomatic BP drop to 62/46 with HR 78 (pre-tilt BP 111/52 with HR 62) at the end of the tilt table when blood was removed from an in situ cannula, with the BP having only a modest transient recovery. UCL4 was only able to tolerate the tilt test for 5.5mins, at which point there was a spontaneous symptomatic fall in BP and HR to 43/32 and 53 respectively (supine BP 126/69 with HR 68). Of note, UCL4 also received a diagnosis of postural orthostatic tachycardiac syndrome (POTS), due to having a sustained HR rise of >30 on supine/standing measures and a HR rise of 32 at 4mins of tilt, associated with symptoms of orthostatic intolerance. UCL5 experienced a spontaneous, symptomatic gradual fall in BP and HR to 66/33 and 57 respectively at 8 minutes of head up tilt (supine 113/74 with HR 59). UCL8 experienced a symptomatic fall in BP and HR to 52/18 and 62 respectively, provoked by venepuncture during head-up tilt (supine 99/63 with HR 76). UCL11 experienced a symptomatic fall in BP and HR to 73/38 and 37 respectively, provoked by venepuncture during head-up tilt (supine 134/75 with HR 42).

One patient (UCL2) received a diagnosis of neurogenic orthostatic hypotension. He experienced a symptomatic BP drop with reflex HR increase during head-up tilt, suggestive of sympathetic vasoconstrictor failure with sparing of parasympathetic cardiac vagal function.

Five additional patients experienced symptoms during head-up tilt although there was no objective significant change in BP or HR. UCL1, UCL7, UCL9, UCL12 reported “dizziness” whilst tilted, and UCL6 had an episode of dissociative collapse whilst tilted.

Five patients did not have any abnormalities on routine autonomic function testing (UCL9, UCL12, UCL13, UCL16, UCL17). One of these (UCL16) was discovered to have an alternative diagnosis for his symptoms when nerve conduction studies showed evidence of central somatosensory pathway damage in the limb experiencing vasomotor symptoms.

### Illustrative case – UCL5

Patient UCL5 was a male in his 40s, who had sustained a moderate-severe TBI in his early 30s. He reported multiple problems typical of patients living with a moderate-severe TBI, such as memory and attention problems, as well as multiple episodes of collapse with loss of consciousness, usually triggered by postural change, which had been occurring since his injury. There were no seizure markers and his previous epilepsy investigations had concluded that this was not the cause of his collapses. He had normal echocardiogram and 24hr Holter, and was being considered for a Reveal device. He had no other medical history, and not on medication. He had autonomic function tests, including a prolonged tilt. He had a normal tilt table assessment and response to cold pressors. However, both his VR and HR_DB_ was below the range for his age (VR=1.26, HR_DB_ =9)^12^, consistent with mild parasympathetic impairment. It was concluded at the time that he had no overall evidence of autonomic dysfunction. However, his symptoms persisted and he underwent follow-up autonomic function tests 2 years later. Testing confirmed the evidence of further worsening of HR variability (VR=1.38, HR_DB_ = 7), and he also experienced a symptomatic drop in BP after 8 minutes of head up tilt. Given the history of recurrent vasovagal syncope episodes,, also reproduced on formal autonomic testing, the patient was referred to our syncope clinic for further management.

## DISCUSSION

We investigated autonomic dysfunction after moderate-severe TBI (msTBI) in two separate patient cohorts with sex and age constitution typically seen in TBI. In a prospectively identified cohort, we found evidence for multi-domain autonomic symptom burden. Furthermore, we show, using a retrospectively identified cohort, that objective autonomic dysfunction is also seen after TBI, and comprises deficits in both sympathetic and parasympathetic function.

Whilst the presence and impact of cognitive and psychiatric symptoms is increasingly recognised in patients with chronic msTBI, there is less recognition of the systemic effects of TBI. Although somatic symptoms are widely reported^1^, there has been little investigation into their cause or consequence. In the acute setting, autonomic dysfunction presenting as paroxysmal sympathetic hyperactivity is well documented after msTBI, and also has been repeatedly shown to correlate with worse mid-long term functional outcomes^7^. There is some suggestion, from studies assessing heart rate variability in small groups of TBI patients, that cardiovascular autonomic dysfunction persists in chronic TBI, and can manifest as clinically relevant symptoms^2–4^. Our retrospective cohort presented with mild autonomic cardiovascular dysfunction within the spectrum of intermittent autonomic dysfunction, and in particular either new onset vasovagal syncope or postural tachycardia syndrome. One patient had confirmed neurogenic orthostatic hypotension with onset post TBI in the absence of other underlying pathology which might affect the autonomic nervous system. Patients may also have had impairment in other domains including pupillary, sudomotor and gastrointestinalm which were not formally investigated in our retrospective cohort. Our study adds to the current literature by demonstrating that objective and clinically relevant autonomic dysfunction is observed after msTBI. We show this in msTBI patients who are many months or years after their initial injury, and assumed to be ‘fully recovered’.

Our results have important clinical implications. We demonstrate that formal autonomic testing, repeated if necessary, can be extremely helpful for providing diagnostic clarity in this cohort, especially if there are persistent unexplained symptoms. This enables both appropriate management of the patient’s symptoms and the avoidance of further diagnostic burden. Intermittent autonomic dysfunction, such as vasovagal syncope, is treatable. Identifying these in the early stage of the disease is likely to improve long-term quality of life in these patients. Neurogenic orthostatic hypotension is a frequent cause of fall and autonomic testing provide information regarding degree of impairment and guide treatment.

It is interesting to note that the retrospective cohort were largely referred for orthostatic symptoms, whereas the symptoms reported by patients in the prospective cohort included a high burden of other types of symptoms, notably pupillomotor (i.e. sensitivity to light and trouble focussing) and gastrointestinal (i.e. bloating and nausea after meals, constipation and diarrhoea) symptoms. This may reflect the fact that postural symptoms are more readily recognised by non-specialists as being potentially mediated by autonomic dysfunction, and patients are referred as part of the work-up for collapse. It may also be that symptoms captured by the COMPASS31 questionnaire are partially or fully mediated by other post-TBI mechanisms. For example, it is possible that sensitivity to light is partially mediated by new or increased migraines after TBI. However, the occurrence of these symptoms in patients who did not report other features of migraine, and the multi-domain symptom burden, suggests that this cannot be wholly the case. Indeed, it is just as possible that some symptoms, just as light-sensitivity and nausea, are considered non-specific and misattributed to simply “recovering from a bad trauma”. For example, the high burden of gastrointestinal symptoms may reflect damage to central autonomic pathways, such as the motor nuclei of the vagus nerve, as a result of brainstem white matter damage sustained during TBI. This mechanism may be underappreciated by non-autonomic specialists, leading to missed opportunities to investigate appropriately. Indeed, the majority of patients referred for autonomic testing reported experiencing symptoms ‘ever since their injury’, but were not referred until years later. Therefore, it is highly likely that autonomic symptoms are under-recognised, and referrals not made to specialist clinics for further investigation and management.

The frequency of gastrointestinal symptoms is also interesting in the context of recent animal work and small-scale clinical reports, which have found that gastrointestinal permeability and inflammation is also observed after TBI, which can in turn exacerbate central nervous system inflammation and also result in autonomic nervous system dysfunction^16–19^. Given that the mechanisms by which TBI leads to these gastrointestinal effects are not fully known, it is possible that acute autonomic dysfunction contributes to this. Alternatively, gastrointestinal symptoms may be a biomarker for presence or severity of autonomic dysfunction in other domains, or reflect non-autonomic injury-mediated gastrointestinal dysfunction. Further animal and human studies are required to investigate these questions.

Autonomic dysfunction after TBI may occur through a number of mechanisms. White matter injury is considered the hallmark injury of moderate-severe TBI, occurring at the time of injury due to axonal shearing and continuing due to inflammation and delayed axonal degeneration in the chronic period, and resulting in network disruption^20,21^. Injury may have occurred to regions of the central autonomic network (CAN^8^) or, more pertinently in TBI, to white matter tracts linking these regions. Brainstem nuclei and white matter connections emanating to and from thalamic and basal ganglia regions may be particularly vulnerable to damage, which is also linked to catecholaminergic dysfunction contributing to cognitive dysfunction post-TBI^22–24^. Therefore, given the importance of brainstem, thalamic and basal ganglia circuits to autonomic function, injury to these white matter tracts may lead to centrally-mediated autonomic dysfunction after TBI. An indirect way in which TBI can lead to autonomic dysfunction is through an interaction with an underlying vulnerability, for example, a tendency towards hypermobility. Another important indirect mechanism by which TBI can result in autonomic dysfunction is by increasing the risk of conditions which are independent risk factors for autonomic dysfunction, for example neurodegenerative disease^25^ and alcohol abuse^26,27^.

The consequences of autonomic dysfunction for TBI patients are potentially profound and far-reaching. Compared to their counterparts without autonomic dysfunction, worse quality of life and psychological distress is reported in otherwise healthy patients with vasovagal syncope or POTS, and in neurological patients with concurrent autonomic dysfunction^28–30^. Additionally, there is significant overlap in regions of the brain important for autonomic function and those important in cognition and emotional processing, for example, hypothalamus, amygdala and anterior cingulate cortex^31,32^, which means that autonomic dysfunction may be an important biomarker for cognitive/psychological dysfunction. Interoception, the “signalling and perception of internal bodily sensations” is important for both cognitive and emotional function^33,34^. If bodily responses to cognitive or emotional stimuli are compromised as a result of autonomic dysfunction after TBI^4^, then autonomic dysfunction may be an important contributor to cognitive and psychological morbidity after TBI. More broadly, post-TBI disruption to vagal-splenic interactions as a result of central autonomic dysfunction may contribute to observed ongoing systemic inflammation in the chronic stage of TBI^35,36^.

### Limitations

The autonomic function testing cohort was retrospectively recruited, which makes it vulnerable to referral bias (as described above) and potential confounds. However, we excluded comorbidities which have high risk of independently causing autonomic dysfunction. On the other hand, TBI itself may increase the risk or cause these comorbidities, or lead to autonomic dysfunction and the comorbidity in parallel. The small numbers in the autonomic testing cohort mean that we cannot make a comment on prevalence of objective autonomic dysfunction after TBI, though the prospective symptom cohort suggests that it is a substantial proportion.

Although the COMPASS31 has been used in multiple populations with definite and possible autonomic dysfunction, it has not been validated in TBI. The orthostatic symptoms are weighted very highly, whereas many TBI patients reported gut and bladder symptoms, so it is possible that the COMPASS31 misses some autonomic symptoms in this cohort. Future studies would benefit from additional, more focussed symptom assessment in non-orthostatic domains, as well as combining both autonomic testing and COMPASS31 assessment.

For largely practical reasons, we only included moderate-severe TBI in our cohorts, so our findings may not be generalisable to mild/repetitive TBI. Autonomic dysfunction has been reported in mild TBI^38^ so it would be interesting to include such patients in future studies, though the mechanisms through which mild TBI leads to deficits is likely to be different than in moderate-severe TBI.

## CONCLUSION

We present evidence for clinically relevant, subjective and objective, autonomic dysfunction after moderate-severe TBI. We hope these findings will increase awareness of autonomic dysfunction after TBI, how this may underlie systemic symptoms, and promote more diagnostic engagement with these symptoms. Future work should seek to further systematically characterise the nature of autonomic dysfunction, to understand the mechanisms by which TBI leads to these problems, and the links between autonomic dysfunction and other post-TBI sequelae.

## Data Availability

Raw scores from the COMPASS31 questionnaire and clinical autonomic function testing reports from the retrospective cohort are available on request to the corresponding author.

## REFERENCES

1. Ruet, A. et al. A detailed overview of long-term outcomes in severe traumatic brain injury eight years post-injury. Frontiers in Neurology 10, (2019).

2. Grubb, B. P., Kanjwal, K., Karabin, B. & Kanjwal, Y. Autonomic dysfunction presenting as postural tachycardia syndrome following traumatic brain injury. Cardiology Journal 17, 482–487 (2010).

3. Hilz, M. J. et al. Severity of traumatic brain injury correlates with long-term cardiovascular autonomic dysfunction. Journal of Neurology 264, 1956–1967 (2017).

4. Amorapanth, P. X. et al. Traumatic brain injury results in altered physiologic, but not subjective responses to emotional stimuli. Brain Injury 32, 1712–1719 (2018).

5. Pertab, J. L. et al. Concussion and the autonomic nervous system: An introduction to the field and the results of a systematic review. NeuroRehabilitation 42, (2018).

6. Purkayastha, S., Stokes, M. & Bell, K. R. Autonomic nervous system dysfunction in mild traumatic brain injury: a review of related pathophysiology and symptoms. Brain Injury 33, (2019).

7. Meyfroidt, G., Baguley, I. J. & Menon, D. K. Paroxysmal sympathetic hyperactivity: the storm after acute brain injury. The Lancet Neurology 16, (2017).

8. Benarroch, E. E. Physiology and Pathophysiology of the Autonomic Nervous System. CONTINUUM: Lifelong Learning in Neurology 26, (2020).

9. Sletten, D. M., Suarez, G. A., Low, P. A., Mandrekar, J. & Singer, W. COMPASS 31: A Refined and Abbreviated Composite Autonomic Symptom Score. Mayo Clinic Proceedings 87, (2012).

10. Low, P. A., Tomalia, V. A. & Park, K.-J. Autonomic Function Tests: Some Clinical Applications. Journal of Clinical Neurology 9, (2013).

11. Malec, J. F. et al. The Mayo Classification System for Traumatic Brain Injury. Journal of Neurotrauma 1424, 1417–1424 (2007).

12. Autonomic Failure. vol. 1 (Oxford University Press, 2013).

13. Koay, S. et al. Multimodal Biomarkers Quantify Recovery in Autoimmune Autonomic Ganglionopathy. Annals of Neurology 89, (2021).

14. D’Amato, C. et al. The diagnostic usefulness of the combined COMPASS 31 questionnaire and electrochemical skin conductance for diabetic cardiovascular autonomic neuropathy and diabetic polyneuropathy. Journal of the Peripheral Nervous System 25, (2020).

15. Kim, Y. et al. The composite autonomic symptom scale 31 is a useful screening tool for patients with Parkinsonism. PLOS ONE 12, (2017).

16. Hanscom, M., Loane, D. J. & Shea-Donohue, T. Brain-gut axis dysfunction in the pathogenesis of traumatic brain injury. Journal of Clinical Investigation 131, e143777–undefined (2021).

17. Hanscom, M. et al. Acute colitis during chronic experimental traumatic brain injury in mice induces dysautonomia and persistent extraintestinal, systemic, and CNS inflammation with exacerbated neurological deficits. Journal of Neuroinflammation 18, (2021).

18. Olsen, A. B. et al. Effects of traumatic brain injury on intestinal contractility. Neurogastroenterology & Motility 25, (2013).

19. Tan, M., Zhu, J.-C. & Yin, H.-H. Enteral nutrition in patients with severe traumatic brain injury: reasons for intolerance and medical management. British Journal of Neurosurgery 25, (2011).

20. Johnson, V. E., Stewart, W. & Smith, D. H. Axonal pathology in traumatic brain injury. Exp Neurol 246, 35–43 (2013).

21. Hill, C. S., Coleman, M. P. & Menon, D. K. Traumatic Axonal Injury: Mechanisms and Translational Opportunities. Trends Neurosci 39, 311–324 (2016).

22. Jenkins, P. O., Mehta, M. A. & Sharp, D. J. Catecholamines and cognition after traumatic brain injury. Brain 139, 2345–2371 (2016).

23. Simoni, S. de et al. Altered caudate connectivity is associated with executive dysfunction after traumatic brain injury. Brain 141, 148–164 (2018).

24. Jenkins, P. O. et al. Dopaminergic abnormalities following traumatic brain injury. Brain 141, 797–810 (2018).

25. Wilson, L. et al. The chronic and evolving neurological consequences of traumatic brain injury. The Lancet Neurology 16, (2017).

26. Julian, T. H., Syeed, R., Glascow, N. & Zis, P. Alcohol-induced autonomic dysfunction: a systematic review. Clinical Autonomic Research 30, (2020).

27. Weil, Z. M., Corrigan, J. D. & Karelina, K. Alcohol Use Disorder and Traumatic Brain Injury. Alcohol Research 39, 171–180 (2018).

28. Benrud-Larson, L. M. et al. Quality of Life in Patients With Postural Tachycardia Syndrome. Mayo Clinic Proceedings 77, (2002).

29. Tomic, S., Rajkovaca, I., Pekic, V., Salha, T. & Misevic, S. Impact of autonomic dysfunctions on the quality of life in Parkinson’s disease patients. Acta Neurologica Belgica 117, (2017).

30. Ng, J., Sheldon, R. S., Ritchie, D., Raj, V. & Raj, S. R. Reduced quality of life and greater psychological distress in vasovagal syncope patients compared to healthy individuals. Pacing and Clinical Electrophysiology (2018) doi:10.1111/pace.13559.

31. Critchley, H. D. et al. Human cingulate cortex and autonomic control: converging neuroimaging and clinical evidence. Brain 126, (2003).

32. Neudorfer, C. et al. Mapping autonomic, mood, and cognitive effects of hypothalamic region deep brain stimulation. Brain (2021) doi:10.1093/brain/awab170.

33. Kreibig, S. D. Autonomic nervous system activity in emotion: A review. Biological Psychology 84, (2010).

34. Garfinkel, S. N., Seth, A. K., Barrett, A. B., Suzuki, K. & Critchley, H. D. Knowing your own heart: Distinguishing interoceptive accuracy from interoceptive awareness. Biological Psychology 104, (2015).

35. Kumar, R. G., Boles, J. A. & Wagner, A. K. Chronic Inflammation After Severe Traumatic Brain Injury. Journal of Head Trauma Rehabilitation 30, (2015).

36. Pavlov, V. A. & Tracey, K. J. The vagus nerve and the inflammatory reflex—linking immunity and metabolism. Nature Reviews Endocrinology 8, (2012).

37. Novak, P. Orthostatic Cerebral Hypoperfusion Syndrome. Frontiers in Aging Neuroscience 8, (2016).

38. Esterov, D. & Greenwald, B. Autonomic Dysfunction after Mild Traumatic Brain Injury. Brain Sciences 7, (2017).

